# An Information-Theoretic Perspective on Multi-LLM Uncertainty Estimation

**DOI:** 10.1101/2025.07.09.25331207

**Authors:** Maya Kruse, Majid Afshar, Saksham Khatwani, Anoop Mayampurath, Guanhua Chen, Yanjun Gao

**Affiliations:** University of Colorado Anschutz Medical Campus; University of Colorado Boulder; University of Wisconsin Madison

## Abstract

Large language models (LLMs) often behave inconsistently across inputs, indicating uncertainty and motivating the need for its quantification in high-stakes settings. Prior work on calibration and uncertainty quantification often focuses on individual models, overlooking the potential of model diversity. We hypothesize that LLMs make complementary predictions due to differences in training and the Zipfian nature of language, and that aggregating their outputs leads to more reliable uncertainty estimates. To leverage this, we propose MUSE (Multi-LLM Uncertainty via Subset Ensembles), a simple information-theoretic method that uses Jensen-Shannon Divergence to identify and aggregate well-calibrated subsets of LLMs. Experiments on binary prediction tasks demonstrate improved calibration and predictive performance compared to single-model and naïve ensemble baselines.

## 1 Introduction

Although large language models (LLMs) have shown remarkable performance in a wide range of NLP tasks and domains, their output is not always consistent or reliable (Xiao et al., 2022; Zhao et al., 2024b). The same LLM can generate divergent responses under different decoding settings, even with identical inputs (Wang et al., 2024a; Wei et al., 2022). As LLMs enter high-stakes domains like healthcare, quantifying output variance is essential for trust, safety, and decision-making (Gao et al., 2024b; Savage et al., 2025; Qin et al., 2024). Quantifying uncertainty is essential to address this challenge: Generating responses with appropriately calibrated confidence helps determine when the answer is trustworthy (Geng et al., 2024). Although prior work has explored uncertainty estimation and calibration through sampling and self-consistency (Rivera et al., 2024; Gao et al., 2024a; Ling et al., 2024), uncertainty-aware training (Liu et al., 2024; Chen and Mueller, 2024; Kapoor et al., 2024), reflection (Zhao et al., 2024a; Zhang et al., 2024b), ranking (Huang et al., 2024), and conformal prediction (Wang et al., 2024b), these methods focus on single LLMs.

This paper introduces a novel approach to uncertainty quantification by aggregating predictions from multiple LLMs. Different LLMs generalize better in distinct regions of the input space, due to the Zipfian nature of language and differences in training corpora, objectives, and architectures (Piantadosi, 2014; Chan et al., 2022). Based on this, we *hypothesize that* combining their outputs offers a principled way to reduce uncertainty, improve robustness, and better approximate ground truth in regions where individual models may falter.

Specifically, we formulate the problem through an information-theoretic lens, using JensenShannon Divergence (JSD) to capture the degree of disagreement among models. JSD offers a symmetric and bounded measure of divergence between probability distributions (Cover, 1999), making it well-suited for comparing predictions across multiple LLMs. We quantify model disagreement to identify reliable consensus and propose Muse (**M**ulti-LLM **U**ncertainty via **S**ubset **E**nsembles), a simple algorithm that selects and aggregates LLM outputs to balance diversity and reliability.

We evaluate Muse on three publicly available binary prediction datasets. *TruthfulQA* (TQA) covers general domain questions and answers designed to investigate the truthfulness of the model (Lin et al., 2022); both *EHRShot* (Wornow et al., 2023), and *MIMIC-Extract* (MIMIC) are structured clinical datasets derived from records of hospitalized patients in the real world (Johnson et al., 2016; Wang et al., 2020). Focusing on binary prediction enables straightforward evaluation of both discrimination and calibration, and empirical findings provide evidence to support our hypothesis with Muse improving calibration and robustness.

## 2 Related Work

In addition to the related work discussed in §1, Ling et al. (2024) estimate both aleatoric and epistemic uncertainty using entropy within single-LLM incontext learning. Chen et al. (2025) focus on clinical prediction tasks, applying deep ensembles and Monte Carlo dropout to capture uncertainty from a single decoder. While these methods operate within a single-model setting, our work addresses uncertainty in a multi-LLM context. In this space, Zhang et al. (2024a) quantify uncertainty across LLMs via semantic similarity in long-form generation, and Dey et al. (2025) select LLMs from a pool to reduce hallucinations based on task accuracy. In contrast, we propose an information-theoretic framework that selects LLM subsets by minimizing predictive uncertainty via JSD and entropy.

## 3 LLM Uncertainty Quantification

We establish two methods for uncertainty quantification for a single LLM as: (1) *self-consistency-based empirical estimation*, which forms the core of our proposed methods, and (2) *sequence likelihood scoring*, used as a widely adopted baseline in prior work (Geng et al., 2024). **(1) Self-Consistency with Empirical Frequency**. Given a binary classification input, we perform stochastic decoding runs *k* in LLM text generation (GEN), using temperature *T* sampling (*T* = 0.7 and *k* = 10), resulting in a set of outputs 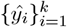. Each output is mapped to a binary label (`yes` or `no`). Define the empirical probability of the label `yes` as:

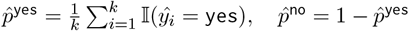

To estimate uncertainty, we apply a bootstrapping procedure: we resample 90% of the outputs with replacement and recompute 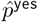 over *B* = 100 trials (denoting as GEN^*BS*^). From the resulting 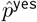 distribution, we compute variance, entropy, and JSD for our proposed algorithms (§ 4). **(2) Sequence Likelihood Scoring**. We adopt the sequence likelihood (SLL) approach used in prior LLM calibration work. For each input *x*, we compute the total log-likelihood of two candidate completions: `“Answer is Yes”` and `“Answer is No”`, denoted LL_yes_ and LL_no_. These are computed using left-to-right autoregressive decoding: 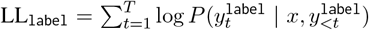. The final prediction probability is obtained via softmax normalization. We use this predicted distribution to compute metrics such as AUROC, ECE and Brier Scores (Guo et al., 2017). Although not robust to

### Algorithm 1

Muse-Greedy version

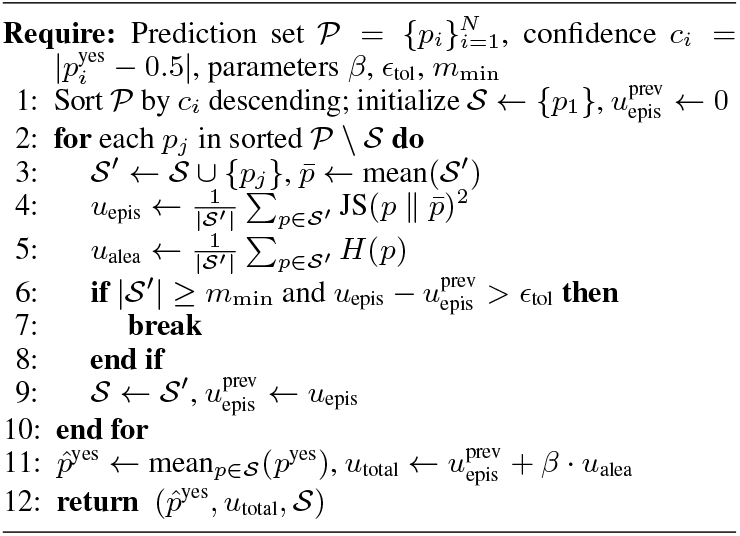

LLM output variability, SLL provides a deterministic scoring baseline for comparison.

## 4 Multi-LLM selective algorithm

When multiple LLMs generate predictive distributions over binary outcomes, we interpret the disagreement among these distributions as a signal of *epistemic uncertainty*, which arises from incomplete knowledge, while consensus suggests more reliable generalization. This can be measured by JSD (Cover, 1999). Meanwhile, we compute the mean entropy *H* of individual model predictions to reflect *aleatoric uncertainty*, capturing inherent input ambiguity. By a novel algorithm that identifies subsets of models 𝒮 whose predictions exhibit low disagreement and low intrinsic uncertainty, this formulation enables us to surface high-consensus regions of the input space while balancing the trade-off between diversity (which may include useful signals) and noise (which can degrade calibration and accuracy).

### Problem Setup

Given an input *x*, let 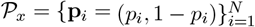 be *N* predictive distributions from multiple LLMs and/or decoding runs, where *p*_*i*_ denotes the predicted probability of the label `yes`. Our goal is to select a subset 𝒮_*x*_ ⊆ *𝒫*_*x*_ that yields a well-calibrated, aggregated prediction 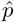.

### Uncertainty Computation

The two types of uncertainty plays an important role in the proposed algorithm. Epistemic uncertainty 𝒰_epis_(𝒮 reflects inter-model disagreement and is quantified as the average JSD between each prediction **p**_*i*_ and the subset mean 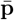:

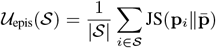

Aleatoric uncertainty 𝒰_alea_(𝒮) reflects intrinsic noise and is estimated by the average binary entropy:

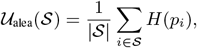

where *H*(*p*) = −*p* log *p* − (1 − *p*) log(1 − *p*). We focus on *optimizing epistemic uncertainty*, as aleatoric uncertainty stems from inherent data noise and is not reducible via model selection. The total uncertainty of a subset of LLMs, denoted as 𝒮, is defined as the sum of its epistemic and aleatoric components: *U* (𝒮) = 𝒰_epis_(𝒮)+*β 𝒰* _alea_(𝒮), where *β* is a weighting factor that controls the trade-off between epistemic disagreement and inherent input ambiguity. Results using total uncertainty *U* (𝒮) are reported in Appendix A.2.

### Multi-LLM Uncertainty via Subset Ensemble

The key contribution of this paper is MUSE, an algorithm that constructs well-calibrated ensembles of LLMs output based on 𝒰_epis_(𝒮). It supports two subset selection strategies, *greedy* and *conservative*, which incrementally select a subset of LLMs whose outputs are mutually diverse yet coherent, as determined by pairwise JSD. Two key parameters control the behavior of Muse: the noise threshold (*ϵ*_tol_) and the minimum subset size *m*_*min*_ as diversity constraint, controlling the balance between ensemble breadth and agreement. The **greedy** version starts with the most confident LLM prediction and iteratively adds models that increase the overall 𝒰_epis_(𝒮) (diversity) of the subset, up to a specified tolerance (as in Algo 1.). The **conservative** version, in contrast, selects models that minimize a joint objective combining epistemic and aleatoric uncertainty. This approach encourages diversity while avoiding instability, resulting in a more calibrated and robust ensemble (see Algo.2).

Once a subset is selected, we compute the final predicted probability by averaging the individual LLM predictions within the subset. Two aggregation strategies are deployed: (1) a simple unweighted mean, and (2) an *aleatoric-aware weighting*, where each LLM’s prediction is 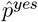 weighted by its 𝒰_alea_(𝒮), where each 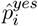 is weighted by its entropy, i.e.,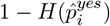. The final prediction is computed as a weighted average, assigning higher weights to more confident (low-entropy) predictions, emphasizing more decisive predictions, particularly when individual models exhibit varying uncertainty levels.

## 5 Experimental Setup

We use *TruthfulQA*(Lin et al., 2022), a benchmark of adversarially designed questions with labeled truthful and untruthful answers. The task is to classify each candidate answer as truthful (`Yes`) or not (`No`), enabling direct evaluation of both discrimination (AUROC) and calibration (ECE, Brier Score). To evaluate performance in a high-stakes setting, we also apply our method to clinical prediction tasks using two structured EHR datasets: diagnosis prediction from *EHRShot*(Wornow et al., 2023) and *MIMIC-Extract* (Wang et al., 2020). On MIMIC, the LLMs predict three outcomes: hospital length of stay ≥ 3 days (LOS3), ≥ 7 days (LOS7), and in-hospital mortality (Mort Hosp.).

We evaluate the following open-source models: Mistral-7B-Instruct (Jiang et al., 2023), Gemma-7B-it (Team et al., 2023), Qwen2-7B-instruct (Yang et al., 2024), and the latest Deepseek-R1-Distil-Qwen-32B (DS-Qwen) (DeepSeek-AI, 2025). All LLMs are run on a server with 4×A100 40GB GPUs. For DS-Qwen, we apply 8-bit quantization to reduce inference time and memory usage. In addition to single LLM method baseline, we compose two naive multi-LLM baselines: majority voting (counting positive labels), and mean of all LLMs’ 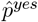 as the final positive probability.

## 6 Results and Discussion

### Overall effectiveness

Muse improves both AU-ROC and calibration metrics compared to single LLMs and naive ensembling baselines, demonstrating its effectiveness in producing reliable and well-calibrated predictions through selective multimodel aggregation. Although the SLL method occasionally yields the highest AUROC, such as DS-Qwen achieving 72.89 on *TruthfulQA*, it often suffers from poor calibration (ECE 57.30, Brier 54.16). In contrast, Muse offers more balanced predictions, with comparable AUROC (72.35) and substantially lower calibration error (ECE 38.15). Similar gains are observed on the EHRShot *acute mi* task, where DS+Mistral+Qwen achieves the best Brier Score (24.4) and strong AUROC (62.2), improving over all single-LLM baselines.

### Balancing diversity and noise

A key strength of our approach is its ability to balance diversity with reliability in multi-LLM ensembles. Figure 1 shows that increasing the minimum subset size (*m*_size_) and moderately relaxing the epistemic uncertainty threshold (*ϵ*_tol_) consistently improves both AUROC and ECE. A larger *m*_size_ (≥ 20) promotes diversity by including more models, while a moderate *ϵ*_tol_ ([0.04, 0.08]) allows controlled disagreement without overwhelming the ensemble with noise. The best performance is achieved when both parameters are carefully balanced. This supports our hypothesis that LLMs offer complementary strengths and provides empirical evidence for our subset-based uncertainty aggregation framework.

**Figure 1.**
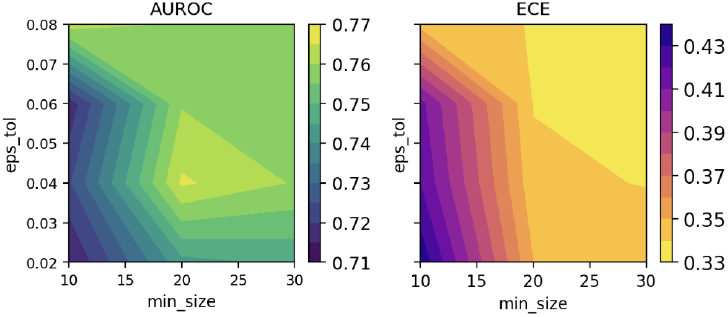
Contour plot of AUROC and ECE as Muse parameters (*m*_*size*_, *ϵ*_tol_) vary, based on a TQA dev set. Main results use *m*_*size*_=20, *ϵ*_tol_=0.04. See Appendix for further analysis.

**Figure 2.**
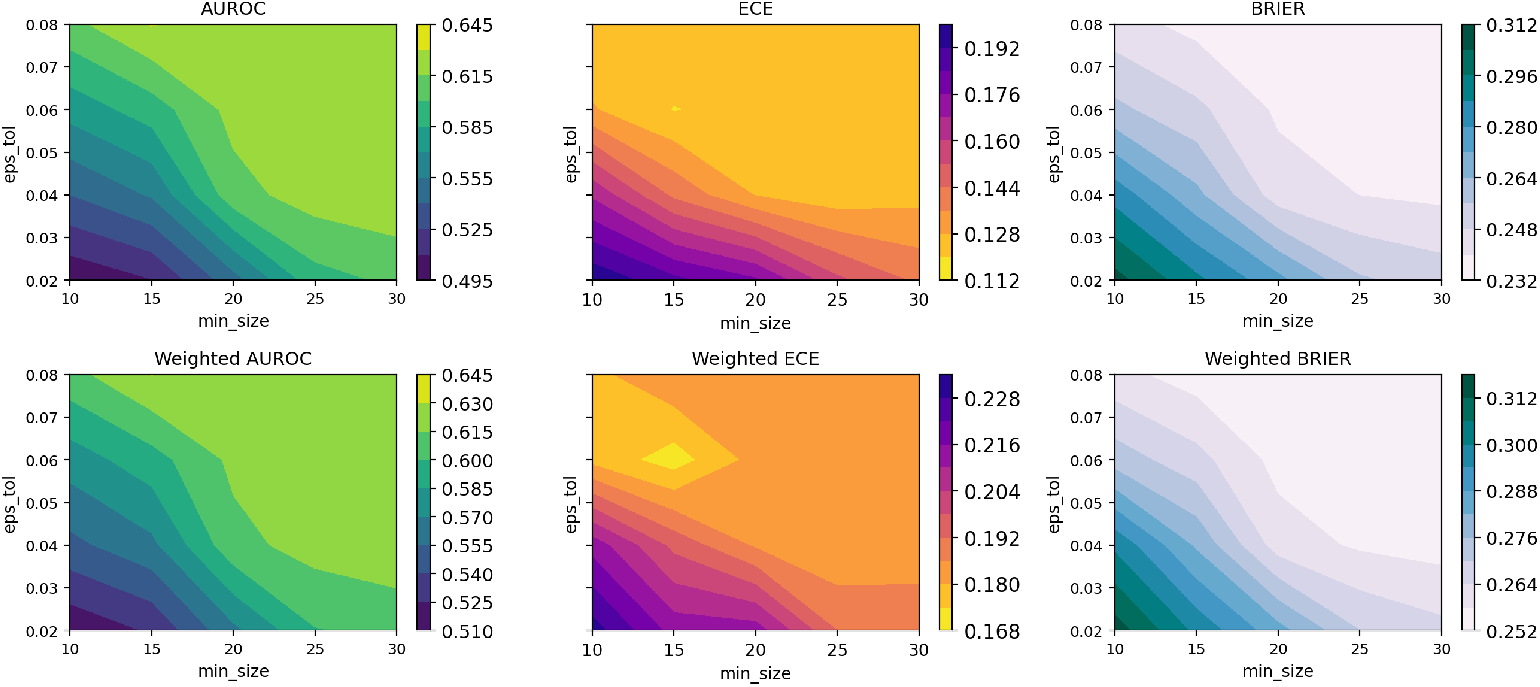
Contour plot for parameter sensitivity analysis using *lupus* prediction task from EHRShot. We report Muse-Greedy with both weighted and unweighted version, to showcase the differences.

### Adaptive model selection

Our method further demonstrates adaptive behavior: performance improves when “stronger” LLMs are present but degrades when weak or noisy models dominate the candidate pool, where strength is defined by each model’s single-task performance. This effect is most evident in *acute mi* and *TQA*. In EHRShot, DS and Mistral form a strong ensemble, but adding a weaker model like Gemma introduces noise that contaminates the pool and harms performance. This supports our hypothesis that selective aggregation for trustable consensus is the key to reliable ensemble performance. Muse makes no assumption that more models lead to better results; rather, it selectively aggregates those that contribute meaningful, calibrated signals to reduce total uncertainty. Table 6 includes a pair of contrasting cases from two prediction tasks in EHRShot for readers who’s interested in (*“weak models dominate”* vs. *“encountered strong, stronger”*) behavior.

## 7 Conclusion

We present Muse, a selective multi-LLM method for uncertainty-aware prediction that improves both accuracy and calibration across domains. Future work will explore dynamic selection strategies and scaling to larger LLM collections in real-world applications, where computational and deployment constraints play a central role.

## Data Availability

All data produced in the present study are available upon reasonable request to the authors

## Limitation

Our study evaluates a limited set of open-source LLMs and focuses exclusively on binary prediction tasks, where evaluation of discrimination and calibration is most straightforward. We also assume access to all model outputs during inference, which may not reflect real-time or resource-constrained deployment scenarios. However, our focus is not on maximizing efficiency, but on understanding how model composition and selective aggregation affect uncertainty estimation. Nonetheless, we have provided empirical evidence that Muse consistently improves both accuracy and calibration, highlighting the value of principled multi-model fusion. Future work will extend to more complex prediction settings and explore efficient selection strategies across broader model ecosystems.

## Ethical Consideration

This study uses two publicly available, deidentified clinical datasets (MIMIC-Extract and EHRShot), ensuring no personally identifiable information is accessed or exposed. All models used are open-source LLMs, and no fine-tuning or data logging was performed, eliminating the risk of patient data leakage. While our focus is on evaluating uncertainty and not clinical deployment, we emphasize the need for responsible use of LLMs in sensitive domains. Any generative outputs or predictions from these models should be interpreted with caution, especially in clinical contexts, and subject to domain expert validation prior to real-world application.

## Algorithm 2

Muse-Conservative version

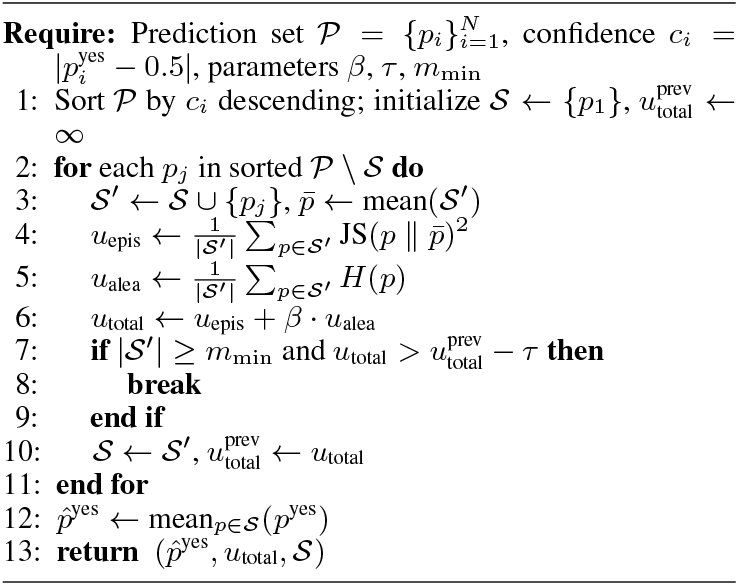

## A More Analysis and Results

### A.1 Muse Conservative Algorithm

Algorithm 2 presents the Muse conservative version. We consider total uncertainty as the sum of epistemic and aleatoric components to better balance diversity and reliability in model selection. The conservative version of Muse adopts a cautious strategy by only adding models when their inclusion leads to a meaningful reduction in total uncertainty. This prevents noisy or unstable predictions from being included, resulting in a more stable and selective ensemble that emphasizes trustworthy aggregation rather than maximizing diversity alone.

### A.2 Comparison of P(Yes) vs. *U* (𝒮)

To compare different scoring strategies, we evaluate the predicted probability *p*(`Yes`) and the total uncertainty (the sum of epistemic and aleatoric components) as predictors of label correctness. As shown in Table 4, *p*(`Yes`) consistently achieves higher AUROC and lower ECE across all tasks, indicating better discrimination and calibration. However, total uncertainty yields lower Brier scores in some cases (e.g., LOS7), suggesting it may better reflect the overall confidence–error trade-off in noisier settings. These results indicate that while *p*(`Yes`) is a strong default for classification, total uncertainty can serve as a complementary signal for soft calibration or abstention.

**Table 1:**
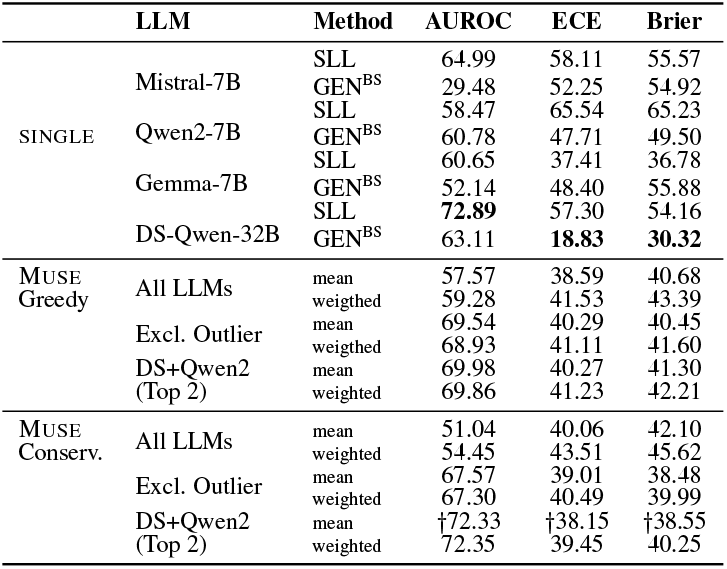
Performance on TruthfulQA. We report SLL and GEN^BS^ results alongside all Muse settings (greedy/conservative, with or without aleatoric weighting). † highlight cases where Muse yields competitive AUROC with lower calibration error despite not being the top performer.

**Table 2:** Performance on the EHRShot Acute myocardial infarction (*acute mi*) task across single LLMs (GEN^BS^)and multi-LLM combinations. Greedy and conservative (nonweighted) results are identical. DS = DS-Qwen.

| LLMs | AUROC ↑ | ECE ↓ | Brier Score ↓ |
| --- | --- | --- | --- |
| Qwen2 | 53.52 | 22.22 | 32.60 |
| Mistral | 60.10 | 13.70 | 25.50 |
| Gemma | 50.20 | **9.80** | 32.30 |
| DS-Qwen | 62.00 | 10.50 | 30.00 |
| DS+Qwen2 | 61.78 | 17.50 | 28.89 |
| DS+Mistr | **64.73** | 16.57 | 25.00 |
| DS+Mistr+Qwen2 | 62.24 | 11.74 | **24.38** |
| All | 60.94 | 12.76 | 24.38 |

**Table 3:**
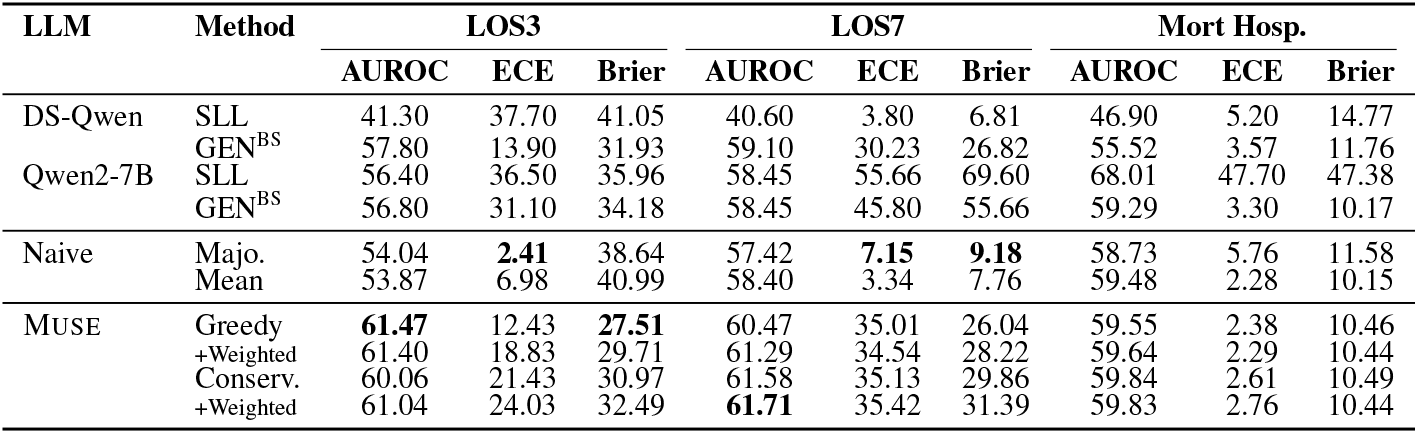
AUROC, ECE, and Brier Score across clinical prediction tasks. We include DS-Qwen and Qwen-7B because they are the two best-performing single LLMs on this dataset (see more results in Table 5). We compare individual LLMs, simple fusion baselines (majority voting “Majo.” and mean), and algorithmic subset selection strategies (greedy and conservative).

**Table 4:** Comparison of predicted probability *p*(Yes) vs. total uncertainty (epistemic + aleatoric) as scoring signals. AUROC favors *p*(`Yes`), while Brier Score occasionally improves with uncertainty-based scoring.

| Metric | P(Yes) | $U(\mathcal{S})$ | Best |
| --- | --- | --- | --- |
| *LOS 3* |  |  |  |
| AUROC | **0.5804** | 0.5103 | P(Yes) |
| ECE | **0.2015** | 0.2208 | P(Yes) |
| Brier Score | 0.3321 | **0.3168** | $U(\mathcal{S})$ |
| *LOS 7* |  |  |  |
| AUROC | **0.5748** | 0.5005 | P(Yes) |
| ECE | **0.1998** | 0.2135 | P(Yes) |
| Brier Score | 0.3514 | **0.1519** | $U(\mathcal{S})$ |
| *Mortality* |  |  |  |
| AUROC | **0.5598** | 0.5263 | P(Yes) |
| ECE | **0.0283** | 0.1140 | P(Yes) |
| Brier Score | **0.1073** | 0.1085 | P(Yes) |

### A.3 More results on MIMIC-Extract

Table 5 presents the performance of two single LLMs and the Muse multi-LLM approach across three clinical prediction tasks using three uncertainty estimation methods: sequence likelihood (SLL), generation-based prediction with bootstrapped generation (GEN^BS^). The results show that Muse, particularly with the Greedy v2 strategy that minimizes total uncertainty, consistently improves AUROC while also reducing calibration error and Brier score compared to individual LLMs. For instance, in the Mortality prediction task, Greedy v2 achieves the highest AUROC (62.88) and the lowest Brier score (9.51), outperforming both Mistral and Gemma models under all methods. Similarly, in LOS3 and LOS7, Muse achieves competitive or best AUROC while offering substantial improvements in calibration, with ECE as low as 4.02 in Mortality. The Conservative variant further enhances calibration, reaching an ECE of 3.88, though at the cost of slightly lower AUROC. These findings demonstrate the effectiveness of Muse in producing more reliable and better-calibrated predictions by aggregating complementary strengths from multiple LLMs.

**Table 5:** More AUROC, ECE, and Brier Score across clinical prediction tasks (MIMIC-Extract) for Mistral-7B and Gemma-7B. In this table, we also report the Muse results from Mistral, Gemma, DS-Qwen and Qwen.

| LLM | Method | LOS3 |  |  | LOS7 |  |  | Mort Hosp. |  |  |
| --- | --- | --- | --- | --- | --- | --- | --- | --- | --- | --- |
|  |  | AUROC | ECE | Brier | AUROC | ECE | Brier | AUROC | ECE | Brier |
| Mistral-7B | SLL | 41.41 | 11.34 | 26.21 | 46.08 | 44.59 | 27.02 | 44.15 | 28.78 | 18.81 |
|  | GEN<sup>BS</sup> | 51.28 | 15.63 | 28.37 | 55.49 | 38.96 | 25.38 | 52.70 | 14.59 | 14.07 |
| Gemma-7B | SLL | 46.48 | 24.71 | 33.85 | 51.37 | 16.04 | 13.44 | 58.89 | 10.52 | 10.48 |
|  | GEN<sup>BS</sup> | 55.09 | 36.91 | 42.78 | 53.93 | 38.29 | 36.70 | 55.94 | 10.40 | 15.45 |
| MUSE | Greedy | **60.70** | 13.34 | 26.42 | 60.70 | 36.43 | 24.30 | 62.25 | 4.36 | 9.59 |
|  | weighted | 60.24 | 18.38 | 28.34 | **61.02** | 35.14 | 26.07 | **62.88** | 4.02 | **9.51** |
|  | Conserv. | 58.34 | 30.87 | 36.17 | 58.74 | 36.29 | 32.15 | 61.89 | **3.88** | 10.90 |

### A.4 Analyzing the adapting behavior of Muse method via EHRShot

Table 6 illustrates the adaptive behavior of our multi-LLM calibration algorithm. In the hyperlipidemia task, where all individual LLMs perform modestly, the aggregated model underperforms, indicating that combining weak predictors can degrade performance. In contrast, for the lupus task, where strong base models (e.g., Deepseek-Distill) are available, the algorithm adapts effectively, matching the best AUROC while maintaining good calibration. This contrast demonstrates the algorithm’s adaptive strength: it amplifies strong signals when present, but cannot compensate when no reliable model exists.

**Table 6:** Performance comparison across two EHRShot tasks. In *hyperlipidemia*, where weak models dominate, the multi-LLM algorithm underperforms. In *lupus*, encountering stronger base models allows the algorithm to adapt and perform competitively, reflecting the adaptive behavior pattern.

| LLMs | AUROC ↑ | ECE ↓ | Brier Score ↓ |
| --- | --- | --- | --- |
| *Hyperlipidemia (weak models dominate)* |  |  |  |
| Qwen | 46.91 | 38.92 | 50.88 |
| Mistral | 52.53 | 14.47 | 25.69 |
| Gemma | 45.18 | 25.72 | 35.36 |
| Deepseek-Distill | 43.92 | 22.46 | 40.97 |
| Greedy (mean) | 33.16 | 24.51 | 29.13 |
| (weighted) | 33.17 | 27.24 | 31.16 |
| *Lupus (encountered strong, stronger)* |  |  |  |
| Qwen | 45.36 | 53.09 | 47.03 |
| Mistral | 33.89 | 19.38 | 11.39 |
| Gemma | 51.60 | 13.97 | 11.54 |
| Deepseek-Distill | 50.94 | 3.70 | 6.84 |
| Greedy (mean) | 50.56 | 10.94 | 22.48 |
| (weighted) | 52.45 | 11.82 | 23.09 |

**Table 7:** Performance on the EHRShot acute mi task across different multi-LLM combinations. **AUROC Avg. Score** column is a simple average of the bootstrapped score of individual LLMs. **AUROC Algo. Score** signifies the (non-weighted) AUROC score of the combination of LLMs using Greedy approach.

| LLMs Combination | AUROC Naive | AUROC MUSE |
| --- | --- | --- |
| Qwen + Mistral | 56.78 | 55.17 |
| Mistral + Gemma | 55.14 | 55.15 |
| Gemma + Qwen | 51.87 | 54.20 |
| Mistral + Gemma + DS | 57.41 | 65.81 |
| Mistral + Gemma + Qwen | 54.60 | 56.47 |
| Qwen + Gemma + DS | 55.23 | 61.05 |

Table 6 shows the impact of different LLM combinations on AUROC for the EHRShot acute MI task. While naive averaging yields modest performance gains, the Muse algorithm substantially boosts AUROC by selectively aggregating informative models. Notably, combinations with higher average AUROC do not always lead to better algorithmic performance: e.g., Qwen+Mistral ranks highest by average but is outperformed by combinations including DeepSeek when selected adaptively. This reinforces that performance is not simply a function of the number of models, but of their individual quality and how well their signals complement one another.

### A.5 Parameter sensitivity on EHRShot

We evaluate how Muse performance varies with the two key hyperparameters: minimum subset size (*m*_size_) and epistemic tolerance (*ϵ*_tol_), using EHRShot as the evaluation dataset (*lupus* prediction). The top row shows unweighted AUROC, ECE, and Brier scores, while the bottom row shows the same metrics when aleatoric uncertainty is used as weighting in aggregation.

Overall, larger *m*_size_ and moderate *ϵ*_tol_ (0.04–0.08) consistently lead to better performance across all metrics. The gains are especially pronounced in calibration (lower ECE and Brier), showing the benefit of including diverse yet coherent model outputs. Aleatoric weighting further improves stability, particularly under looser inclusion criteria. These trends confirm that careful tuning of subset size and disagreement tolerance is key to balancing diversity and reliability in multi-LLM ensembles.

## References

Stephanie Chan, Adam Santoro, Andrew Lampinen, Jane Wang, Aaditya Singh, Pierre Richemond, James McClelland, and Felix Hill. 2022. Data distributional properties drive emergent in-context learning in transformers. Advances in neural information processing systems, 35:18878–18891.

Jiuhai Chen and Jonas Mueller. 2024. Quantifying uncertainty in answers from any language model and enhancing their trustworthiness. In Proceedings of the 62nd Annual Meeting of the Association for Computational Linguistics (Volume 1: Long Papers), pages 5186–5200.

Zizhang Chen, Peizhao Li, Xiaomeng Dong, and Pengyu Hong. 2025. Uncertainty quantification for clinical outcome predictions with (large) language models. In Findings of the Association for Computational Linguistics: NAACL 2025, pages 7512–7523, Albuquerque, New Mexico. Association for Computational Linguistics.

Thomas M Cover. 1999. Elements of information theory. John Wiley & Sons.

DeepSeek-AI. 2025. Deepseek-r1: Incentivizing reasoning capability in llms via reinforcement learning. Preprint, arXiv:2501.12948.

Prasenjit Dey, Srujana Merugu, and Sivaramakrishnan Kaveri. 2025. Uncertainty-aware fusion: An ensemble framework for mitigating hallucinations in large language models. arXiv preprint arXiv:2503.05757.

Xiang Gao, Jiaxin Zhang, Lalla Mouatadid, and Kamalika Das. 2024a. Spuq: Perturbation-based uncertainty quantification for large language models. In Proceedings of the 18th Conference of the European Chapter of the Association for Computational Linguistics (Volume 1: Long Papers), pages 2336–2346.

Yanjun Gao, Skatje Myers, Shan Chen, Dmitriy Dligach, Timothy A Miller, Danielle Bitterman, Guanhua Chen, Anoop Mayampurath, Matthew Churpek, and Majid Afshar. 2024b. Position paper on diagnostic uncertainty estimation from large language models: Next-word probability is not pre-test probability. In GenAI for Health: Potential, Trust and Policy Compliance.

Jiahui Geng, Fengyu Cai, Yuxia Wang, Heinz Koeppl, Preslav Nakov, and Iryna Gurevych. 2024. A survey of confidence estimation and calibration in large language models. In NAACL-HLT.

Chuan Guo, Geoff Pleiss, Yu Sun, and Kilian Q Weinberger. 2017. On calibration of modern neural networks. In International conference on machine learning, pages 1321–1330. PMLR.

Xinmeng Huang, Shuo Li, Mengxin Yu, Matteo Sesia, Hamed Hassani, Insup Lee, Osbert Bastani, and Edgar Dobriban. 2024. Uncertainty in language models: Assessment through rank-calibration. In Proceedings of the 2024 Conference on Empirical Methods in Natural Language Processing, pages 284–312.

Albert Q. Jiang, Alexandre Sablayrolles, Arthur Mensch, Chris Bamford, Devendra Singh Chaplot, Diego de las Casas, Florian Bressand, Gianna Lengyel, Guillaume Lample, Lucile Saulnier, Lélio Renard Lavaud, Marie-Anne Lachaux, Pierre Stock, Teven Le Scao, Thibaut Lavril, Thomas Wang, Timotheé Lacroix and William El Sayed. 2023. Mistral 7b. Preprint, arXiv:2310.06825.

Alistair EW Johnson, Tom J Pollard, Lu Shen, Li-wei H Lehman, Mengling Feng, Mohammad Ghassemi, Benjamin Moody, Peter Szolovits, Leo Anthony Celi, and Roger G Mark. 2016. Mimic-iii, a freely accessible critical care database. Scientific data, 3(1):1–9.

Sanyam Kapoor, Nate Gruver, Manley Roberts, Arka Pal, Samuel Dooley, Micah Goldblum, and Andrew Wilson. 2024. Calibration-tuning: Teaching large language models to know what they don’t know. In Proceedings of the 1st Workshop on Uncertainty-Aware NLP (UncertaiNLP 2024), pages 1–14.

Stephanie Lin, Jacob Hilton, and Owain Evans. 2022. Truthfulqa: Measuring how models mimic human falsehoods. In Proceedings of the 60th Annual Meeting of the Association for Computational Linguistics (Volume 1: Long Papers), pages 3214–3252.

Chen Ling, Xujiang Zhao, Xuchao Zhang, Wei Cheng, Yanchi Liu, Yiyou Sun, Mika Oishi, Takao Osaki, Katsushi Matsuda, Jie Ji, and 1 others. 2024. Uncertainty quantification for in-context learning of large language models. In Proceedings of the 2024 Conference of the North American Chapter of the Association for Computational Linguistics: Human Language Technologies (Volume 1: Long Papers), pages 3357–3370.

Shudong Liu, Zhaocong Li, Xuebo Liu, Runzhe Zhan, Derek Wong, Lidia Chao, and Min Zhang. 2024. Can llms learn uncertainty on their own? expressing uncertainty effectively in a self-training manner. In Proceedings of the 2024 Conference on Empirical Methods in Natural Language Processing, pages 21635–21645.

Steven T Piantadosi. 2014. Zipf’s word frequency law in natural language: A critical review and future directions. Psychonomic bulletin & review, 21:1112– 1130.

Jeremy Qin, Bang Liu, and Quoc Nguyen. 2024. Enhancing healthcare llm trust with atypical presentations recalibration. In Findings of the Association for Computational Linguistics: EMNLP 2024, pages 2520–2537.

Mauricio Rivera, Jean-François Godbout, Reihaneh Rabbany, and Kellin Pelrine. 2024. Combining confidence elicitation and sample-based methods for uncertainty quantification in misinformation mitigation. In Proceedings of the 1st Workshop on Uncertainty-Aware NLP (UncertaiNLP 2024), pages 114–126.

Thomas Savage, John Wang, Robert Gallo, Abdessalem Boukil, Vishwesh Patel, Seyed Amir Ahmad Safavi-Naini, Ali Soroush, and Jonathan H Chen. 2025. Large language model uncertainty proxies: discrimination and calibration for medical diagnosis and treatment. Journal of the American Medical Informatics Association, 32(1):139–149.

Gemini Team, Rohan Anil, Sebastian Borgeaud, Jean-Baptiste Alayrac, Jiahui Yu, Radu Soricut, Johan Schalkwyk, Andrew M Dai, Anja Hauth, Katie Millican, and 1 others. 2023. Gemini: a family of highly capable multimodal models. arXiv preprint arXiv:2312.11805.

Shirly Wang, Matthew BA McDermott, Geeticka Chauhan, Marzyeh Ghassemi, Michael C Hughes, and Tristan Naumann. 2020. Mimic-extract: A data extraction, preprocessing, and representation pipeline for mimic-iii. In Proceedings of the ACM conference on health, inference, and learning, pages 222–235.

Xinglin Wang, Yiwei Li, Shaoxiong Feng, Peiwen Yuan, Boyuan Pan, Heda Wang, Yao Hu, and Kan Li. 2024a. Integrate the essence and eliminate the dross: Finegrained self-consistency for free-form language generation. In Proceedings of the 62nd Annual Meeting of the Association for Computational Linguistics (Volume 1: Long Papers), pages 11782–11794.

Zhiyuan Wang, Jinhao Duan, Lu Cheng, Yue Zhang, Qingni Wang, Xiaoshuang Shi, Kaidi Xu, Heng Tao Shen, and Xiaofeng Zhu. 2024b. Conu: Conformal uncertainty in large language models with correctness coverage guarantees. In Findings of the Association for Computational Linguistics: EMNLP 2024, pages 6886–6898.

Jason Wei, Xuezhi Wang, Dale Schuurmans, Maarten Bosma, Fei Xia, Ed Chi, Quoc V Le, Denny Zhou, and 1 others. 2022. Chain-of-thought prompting elicits reasoning in large language models. Advances in neural information processing systems, 35:24824– 24837.

Michael Wornow, Rahul Thapa, Ethan Steinberg, Jason Fries, and Nigam Shah. 2023. Ehrshot: An ehr benchmark for few-shot evaluation of foundation models. Advances in Neural Information Processing Systems, 36:67125–67137.

Yuxin Xiao, Paul Pu Liang, Umang Bhatt, Willie Neiswanger, Ruslan Salakhutdinov, and Louis-Philippe Morency. 2022. Uncertainty quantification with pre-trained language models: A large-scale empirical analysis. In Findings of the Association for Computational Linguistics: EMNLP 2022, pages 7273–7284.

An Yang, Beichen Zhang, Binyuan Hui, Bofei Gao, Bowen Yu, Chengpeng Li, Dayiheng Liu, Jianhong Tu, Jingren Zhou, Junyang Lin, and 1 others. 2024. Qwen2. 5-math technical report: Toward mathematical expert model via self-improvement. arXiv preprint arXiv:2409.12122.

Caiqi Zhang, Fangyu Liu, Marco Basaldella, and Nigel Collier. 2024a. Luq: Long-text uncertainty quantification for llms. In Proceedings of the 2024 Conference on Empirical Methods in Natural Language Processing, pages 5244–5262.

Wenqi Zhang, Yongliang Shen, Linjuan Wu, Qiuying Peng, Jun Wang, Yueting Zhuang, and Weiming Lu. 2024b. Self-contrast: Better reflection through inconsistent solving perspectives. In Proceedings of the 62nd Annual Meeting of the Association for Computational Linguistics (Volume 1: Long Papers), pages 3602–3622, Bangkok, Thailand. Association for Computational Linguistics.

Xinran Zhao, Hongming Zhang, Xiaoman Pan, Wenlin Yao, Dong Yu, Tongshuang Wu, and Jianshu Chen. 2024a. Fact-and-reflection (far) improves confidence calibration of large language models. In Findings of the Association for Computational Linguistics ACL 2024, pages 8702–8718.

Yukun Zhao, Lingyong Yan, Weiwei Sun, Guoliang Xing, Shuaiqiang Wang, Chong Meng, Zhicong Cheng, Zhaochun Ren, and Dawei Yin. 2024b. Improving the robustness of large language models via consistency alignment. In Proceedings of the 2024 Joint International Conference on Computational Linguistics, Language Resources and Evaluation (LREC-COLING 2024), pages 8931–8941.

